# Morbidity associated with *Schistosoma mansoni* infection in north-eastern Democratic Republic of the Congo

**DOI:** 10.1101/2020.05.23.20108654

**Authors:** Maurice M Nigo, Peter Odermatt, David Wully Nigo, Georgette B. Salieb-Beugelaar, Manuel Battegay, Patrick R Hunziker

## Abstract

**Background:** Controlling morbidity is the main target of schistosomiasis control. Yet only rarely do we assess morbidity linked to *Schistosoma* sp. infection. In the Democratic Republic of Congo (DRC), and particularly in the north-eastern Ituri province, morbidity associated with *Schistosoma mansoni* infection is unknown. For this reason, we aimed to assess intestinal and hepatosplenic morbidity associated with *S. mansoni* infection in Ituri province.

**Methods / Principal Findings:** In 2017, we conducted a cross-sectional study in 13 villages in Ituri province, DRC. *S. mansoni* infection was assessed with a Kato-Katz stool test (2 smears) and a point-of-care circulating cathodic antigen (POC-CCA) test in urine. A questionnaire was used to obtain demographic data and information about experienced intestinal morbidity. Each participant underwent an abdominal ultrasonography examination to diagnose hepatosplenic morbidity. Of the 586 study participants, 76.6% tested positive for *S. mansoni*. Intestinal morbidity, such as abdominal pain (52.7%), diarrhoea (23.4%) and blood in the stool (21.5%) in the previous two weeks, was very frequent. Hepatosplenic morbidity was demonstrated by abnormal liver parenchyma patterns (42.8%), hepatomegaly (26.5%), and splenomegaly (25.3%). Liver pathology (adjusted odds ratio [aOR] 1.20, 95% confidence interval [CI] 1.06–1.37, *P*=0.005) was positively and significantly associated with *S. mansoni* infection. Hepatomegaly (aOR 1.52, 95% confidence interval [CI] 0.99–2.32, *P*=0.053) and splenomegaly (aOR 1.12, 95% CI 0.73–1.72, *P*=0.619) were positively but not significantly associated with *S. mansoni* infection at the individual level. At the village level, *S. mansoni* prevalence was positively associated with the prevalence of hepatomegaly and splenomegaly. Higher *S. mansoni* infection intensities were associated with diarrhoea, blood in the stool, hepatomegaly, splenomegaly and with liver parenchyma, pathology patterns C, D, E and F. Four study participants were diagnosed with ascites and five reported hematemesis.

**Conclusions/Significance:** Our study documents a high burden of intestinal and hepatosplenic morbidity associated with *S. mansoni* infection status in Ituri province. The results call for targeted interventions to address both *S. mansoni* infection and related morbidity.

**Author Summary:** Schistosomiasis caused by *Schistosoma mansoni* is of great public health importance in sub-Saharan Africa. The World Health Organization (WHO) recommends that control efforts aim to reduce morbidity through large scale intervention programmes. However, intestinal and liver morbidity is rarely assessed in such control programmes. Hence, little is known about (i) the magnitude of the intestinal and liver morbidity burden in each community, or about (ii) the morbidity associated with *S. mansoni* infection, specifically. We conducted a (cross-sectional) study in which we assessed intestinal morbidity by questionnaire and liver morbidity by abdominal ultrasonography. Further, we determined the infection status of the study participants using standard diagnostic procedures (Kato-Katz technique and point-of-care cathodic circulating *S. mansoni* antigen [POC-CCA] test in urine). Among 586 study participants, six years and older, from 13 villages in Ituri province, DRC, we observed a high degree of intestinal (e.g. 23.4% with diarrhoea, 21.5% with blood in stool) and hepatosplenic morbidity (e.g. 42.8% with abnormal liver patterns C, D, E, and F, 26.5% with enlarged liver, 25.3% with enlarged spleen). *S. mansoni* infection was associated with liver and spleen enlargement. Likewise, *S. mansoni* infection intensity was linked to diarrhoea, to liver and spleen enlargement and to pathological changes in the liver parenchyma. At village level, we observed that the prevalence of enlarged liver and spleen among patients increased with the prevalence of *S. mansoni* infection. We conclude that the population of Ituri province carries an alarming burden of intestinal, liver and spleen morbidity associated with *S. mansoni* infection. Therefore, a comprehensive control programme to address this infection and disease burden is urgently required.

## Introduction

Schistosomiasis, a chronic helminth infection is caused by trematodes of the genus *Schistosoma and* belongs to the so-called neglected tropical diseases. It continues to be a major global cause of morbidity and mortality [1].

Depending on the species, it may be of genitourinary *(Schistosoma haematobium)* or of intestinal *(Schistosoma mansoni, S. japonicum, S. mekongi, S. intercalatum*, and *S. guineensis)* form. The basis of the disease is the host’s cell-mediated granulomatous immune response to the soluble antigens of the parasite eggs trapped in the tissues [2, 3]. In the intestinal form the adult worm dwell in the portal vein and mesenteric veinlets that drain the intestines and where the female deposits the eggs during her daily migration. Chronic and heavy infections are frequently associated with hepatosplenic and intestinal disease characterized by liver and spleen enlargements and intestine damage. In the liver, the resulting scars may disrupt liver function and obstruct the portal veins, leading to periportal fibrosis (PPF), portal hypertension, subsequently to oesophageal varices and then to hematemesis and melena, and ultimately ascites, the main direct causes of death due to *S. mansoni* infection [4, 5]. In the intestines, inflammation may induce diarrhoea, whereas granulomas may cause polyposis with ulcers and recurrent bleeding. The resulting intestinal clinical manifestations may include abdominal pain, diarrhoea, and the presence of blood in the stool [2, 6]. Chronic schistosomiasis can also lead to anaemia, stunted growth and impairment of cognitive development [7, 8]. Many infected people, even those with considerable infection intensity, may remain asymptomatic for a long period or experience only non-specific symptoms, such as nausea, headaches, fever, fatigue and abdominal pain [9].

Preventive chemotherapy (PCT) mass drug administration is the WHO recommended strategy for both the reduction of morbidity and the control of schistosomiasis in endemic settings [10]. However, for several reasons, this recommendation is poorly implemented in many countries, partly because of lack of commitment, funding and/or political instabilities and security issues.

In the Democratic Republic of Congo (DRC), the full extent of schistosomiasis morbidity burden remains unknown and most of the relevant information about the disease date back for more than twenty years [11, 12]. The existing publications report on *Schistosoma* infection. The few reports related to morbidity mainly concern the province of Maniema in central-eastern region of the country [13, 14]. For the Ituri province, morbidity due *S. mansoni* infection was mentioned in colonial times [15]. Since then, only these data and those of 1970s and 1980s are summarized in available reviews. On one hand, Madinga et al. [12] reported that *S. mansoni* endemic areas were described in Ituri before 1954. The main foci being lying on the left bank of Lake Albert, with prevalence rates ranging from 11.0% to 64.9%. Gillet and Wolfs, on the other hand, reported that there were not local cases in the high hill region, and that prevalence ranged from 2.3% in Aru in the north to 93.7% in a fishing village on the shore of Lake Albert [15]. Both these reviews did not mention the existence of *S. haematobium* and *S. intercalatum* infections in the Ituri province [12, 16]. By investigating the prevalence, intensity, and the relative morbidity of *S. mansoni* infection among Ugandan and Zairian school children aged 5 to 20 years in Aru region, Müller et al. found that prevalence was low to moderately high. About 8.0% of children had heavy infection. Among children, 15.6 to 38.0% had hepatomegaly, while 22.0 to 59.2% were diagnosed with splenomegaly. However, they found that association of organomegaly with *S. mansoni* infection was not significant [17].

To the best of our knowledge, no recent study has been undertaken since then until the national survey conducted in between 2013 and 2015. Results of this survey have not yet been published. However, several studies conducted in neighbouring Uganda show high infection and morbidity and considerable mortality linked to *S. mansoni* infection [3, 5, 18]. The aim of the present study was to assess the morbidity associated with *S. mansoni* infection in Ituri province.

## Materials and Methods

### Ethics statement

This study was approved by the Swiss Ethical Commission (Ref. No. UBE-15/78) and by the University of Kisangani’s Research Ethical Commission, (Ref No: CER/003/GEAK/2016). Research authorization was granted by the Nyankunde Higher Institute of Medical Techniques (Ref No 70/ISTM-N/SGAC/2017), Bunia, DRC. Permission for field work was obtained from the Ituri Provincial Health Division (Ref. 054/433/DPS/IT/06/2016 and Ref. 054/472/DPS/IT/06/2017) and from all relevant health districts. Prior to enrolment, the study objectives and procedures were explained to each participant in the local language and all their questions were answered. Written informed consent was obtained from all study participants aged 15 years and older. Parents or legal guardians signed consent forms for participants aged <14 years. Participants diagnosed with *S. mansoni* were treated with praziquantel (40mg/kg) [19]. All participants received Mebendazole (500mg, single dose, Vermox®) for general deworming, in accordance with the DRC national deworming guidelines.

### Study area

This study was conducted in the Ituri province, north-eastern DRC (geographical coordinates: 1.30°–3.60° latitude and 27.00°–31.40° longitude). Ituri province has an area of about 65,658 km^2^ and is home to 5.2 million inhabitants from five different ethnic groups (Nilo-Hamites, Bantu, Nilotic, Sudanese, and Pygmy). It is divided into 5 territories (counties) and 36 health districts, and is bordered by Lake Albert in the east, while several streams and rivers irrigate the province. These waterways are suitable environments for schistosome’s intermediate host snails. This study involved specifically six health districts that were purposively selected because of their high prevalence of *S. mansoni* infection: Angumu, Bunia, Lolwa, Mandima, Nia-Nia, and Tchomia. From these health district, a total 13 villages were purposively selected. Bunia health district being the biggest with a population of more than 500,000 inhabitants, had the lion’s share with 5 selected villages: Lumumba, Simbilyabo, Kindia, Gupe, Sukisa, and Ngezi. For the other health districts, 2 villages from Angumu, namely Gupe and Ndaru-Muswa. 2 from Lolwa: Mambau and Pekele, 2 from Nia-Nia: Bankoko and Mangenengene, 1 from Mandima: Mandima, and 1 from Tchomia: Kadjugi. The presence of *Schistosoma mansoni* has been widely documented in the province in colonial times. The transmission was considered to occur mainly along Lake Albert’s shores [15]. Both the review of the available literature and the consultation of the provincial NTDs control program did not mention the presence of *S. haematobium* in Ituri province and in our prior work in the area (MNN, unpublished information), we did not find S. haematobium. *Schistosoma intercalatum* is mentioned by few authors. For these reasons, we only concentrated our efforts to study morbidity related to intestinal schistosomiasis (*S. mansoni* and *S. intercalatum*) in the Ituri province [12, 15, 16]. Only a small proportion of the population residing in Bunia city has access to an adequate water supply. Most of the population uses natural water bodies (springs, ponds, streams) as its main water source.

### Study design and population

We conducted a cross-sectional, household-based, in-depth study in the 13 purposely selected villages across six health districts in Ituri province. Two stage sampling procedures were to select both households and individuals for the study. At least 10 households were randomly selected in each village and all individuals aged 6 years and older, women and men, present on the day of the survey were enrolled. Household’s visitors, as well as mentally and terminally ill persons were excluded.

The study incorporated household and individual questionnaires; anthropometric assessments; and parasitological, clinical, and abdominal ultrasonographic examinations.

### Procedures

#### Individual questionnaires

All participants were invited to participate in an interview, conducted using a pre-tested questionnaire. This individual questionnaire focused on demographic, anthropometric, occupational, educational, and religious characteristics, as well as on knowledge, attitude and practices related to *S. mansoni* infection and disease. The questionnaire also helped to assess for signs and symptoms related to schistosomiasis, such as diarrhoea or blood in the stool in the previous two weeks, and a history of hematemesis at any time, at least once.

#### Anthropometric measurements

Participants’ height and weight were measured by a Seca analog bathroom scale and height rod and reported to the nearest half kilogram (0.5 kg) and half centimetre (0.5 cm), respectively. Participants’ body mass index (BMI) was also calculated (weight in kilograms divided by the square of the person’s height in metres, kg/m^2^).

#### Parasitological examination

Participants were asked to provide one faecal sample (approx. 5 grams of morning stool) in a labelled plastic container for testing with the Kato-Katz technique [20]. From each stool specimen, two thick smears of 41.7 mg [20] were prepared and examined by experienced technicians. To allow for hookworm assessment, all smears were examined by microscope within one hour after preparation. All slides were examined for *S. mansoni* within 24 hours after stool collection. One third of the prepared smears were checked by the principal investigator. All helminth eggs were counted and recorded for each species separately. The intensity of the helminth infection was calculated by multiplying the mean number of eggs found on the two slides by 24. The result was expressed as eggs per gram (EPG) of stool [8].

Participants were also asked to provide a urine sample (approx. 60 ml) in a pre-labelled, wide-mouth, plastic container, for the detection of circulating *S. mansoni* antigens using a point-of-care circulating cathodic antigen (POC-CCA) test. Both the stool and urine examinations were performed at the relevant village health centre facility.

The POC-CCA tests were performed according to the manufacturer’s guidelines (Rapid Medical Diagnostics, Pretoria, South Africa). Urine was examined on the day of collection. In cases where the test was postponed until the next day, urine samples were kept in a solar fridge, at 2–8°C (Steca, Germany). Test results were deemed negative if the POC-CCA band did not appear within 20 minutes. Trace, weak, medium, and strong coloured CCA bands were recorded as positive results. Questionable results were discussed among at least two technicians and the principal investigator.

#### Clinical examination

All participants underwent clinical and abdominal ultrasonography examinations. Clinical examinations consisted of physical examinations performed by an experienced physician and assisted by an experienced nurse.

#### Abdominal ultrasound examination

An abdominal ultrasound was performed for each participant, according to Niamey protocol [21] and using a 2.0 MHz convex transducer U-Lite Sonoscanner Ultraportable HD Ultrasound Unit (U-Lite, Sonoscanner, 6, Rue André Voguet, Paris, France). A portable generator (MK, China) and solar powered batteries (for remote villages) were used as electricity sources.

Participants were examined in a supine position. The size of the left lobe from the cranial to the caudal was measured at about 2 cm in the left parasternal line (PSL), from the xyphoid. The height and the width of the spleen were also measured. The length and the width of the spleen were measured, and its texture evaluated. All the measures were taken in centimetres and performed using the callipers on the screen the device according the manufacturer recommendations. Organs’ measures were then adjusted to the height of the individual and compared with those of the Senegalese healthy control individuals [22]. The organs were considered enlarged when the size values exceeded two standard deviations (2 SD) from the mean adjusted values of the reference control individuals. Liver parenchyma patterns (Figure S1) were assessed following the WHO/TDR guidelines as grade A: normal, and B: incipient, C: probable, D, E, and F as frank periportal fibrosis [21]. The inner portal vein diameter was measured. The inner gall bladder length and width, and the wall thickness were also measured.

### Data management and analysis

Data was entered in Excel and cross-checked against the data sheet. STATA, version14.2 software (Stata Corp, College Station, USA) was used for data management and analysis. Only participants with a complete dataset were retained in the analysis (Figure 1). Seven age groups were established: (i) 6–9 years, (ii) 10–14 years, (iii) 15–19 years, (iv) 20–29 years, (v) 30–39 years, (vi) 40–49 years, and ≥50 years. Body mass index (BMI) was calculated and four categories were set: underweight (<18.5 kg/m^2^), normal weight (18.5–24.9 kg/m^2^), overweight (25.0–29.9 kg/m^2^), and obese (≥30 kg/m^2^). Infection prevalence was expressed as the number of *S. mansoni-positive* individuals divided by the total number of participants examined. Infection intensity was estimated based on helminth egg counts per gram of stool (EPG) when examined with the Kato-Katz technique [20]. *S. mansoni* infection intensities were classified as light (1–99 EPG), moderate (100–399 EPG), and heavy (≥400 EPG) [8].

Arithmetic mean infection intensity was calculated. Categorical variables were presented as frequencies and percentages. Pearson’s chi-square (χ^2^) test was used to compare frequency distributions. A univariate logistic regression analysis was carried out to identify associations between *S. mansoni* infection status (outcome) and morbidity indicators (predictors) and/or demographic factors (age, gender). Predictors with a significance level of 20% or less, and age and gender variables were included in the multivariable logistic regression models. Odds ratios (OR), adjusted OR (aOR), and corresponding 95% confidence intervals (95% CI) were calculated. *P-*values <0.05 were considered statistically significant. The diagnostic approach combining results of the Kato-Katz technique and those of POC-CCA was used in this study. As a comparison, the same analysis was repeated for the diagnosis with the Kato-Katz technique and the POC-CCA alone.Ultrasonographic measures of organs were defined as enlarged for left liver lobe and spleen length exceeding 2 SD above the normal reference value. Portal vein diameter was considered as enlarged if exceeding 2 SD above the normal reference value. Liver patterns A and B were considered normal, patterns C and D, mild PPF, while patterns E, and F were recorded as severe PPF.

## RESULTS

### Study population

Data were collected between June and September 2017. We enrolled participants from 13 purposely selected villages across six health districts with an anticipated high prevalence of *S. mansoni* infection. Of the 949 individuals enrolled (Figure 1), 586 completed all study procedures and had a complete dataset, that is, one stool sample examined with two Kato-Katz smears, a urine sample tested with POC-CCA, two completed questionnaires, and a clinical and abdominal ultrasound examination.

Among those with a complete dataset, 342 (58.4%) were females, 330 (56.3%) were under 20 years of age, and 268 (45.7%) were underweight (Table 1). The prevalence of *S. mansoni* was 59.2%, 65.7%, and 76.6% according to Kato-Katz, POC-CCA and combined test results, respectively. Thirty-seven percent, 15.2% and 7.2% of the population had light-, moderate- and heavy-intensity infections, respectively. Infection with soil transmitted helminths (STH) was not common among participants, with only eight participants diagnosed with an STH infection. In contrast, intestinal symptoms were very common, with 52.7%, 23.4% and 21.5%, reporting abdominal pain, diarrhoea, and blood in the stool within the two weeks preceding the survey, respectively. Five participants (0.9%) had experienced hematemesis at least once in his/her life. Abdominal ultrasound examinations revealed that 26.5% of participants had hepatomegaly, 25.3% splenomegaly, and 42.8% had liver pathology; 36.4% had mild PPF (patterns C and D), 6.4% had severe PPF (patterns E and F) and 1.2% presented other liver patterns not linked with schistosomiasis (patterns X, Y, and Z), and 0.7% had ascites. Only 56.0% of the participants had a normal liver parenchyma (patterns A and B). More details on liver parenchyma patterns are shown in Table S7 and S8.

**Figure 1:** Flowchart of participant inclusion/exclusion in the 2017 Ituri morbidity study across 13 villages.

**Table 1:**
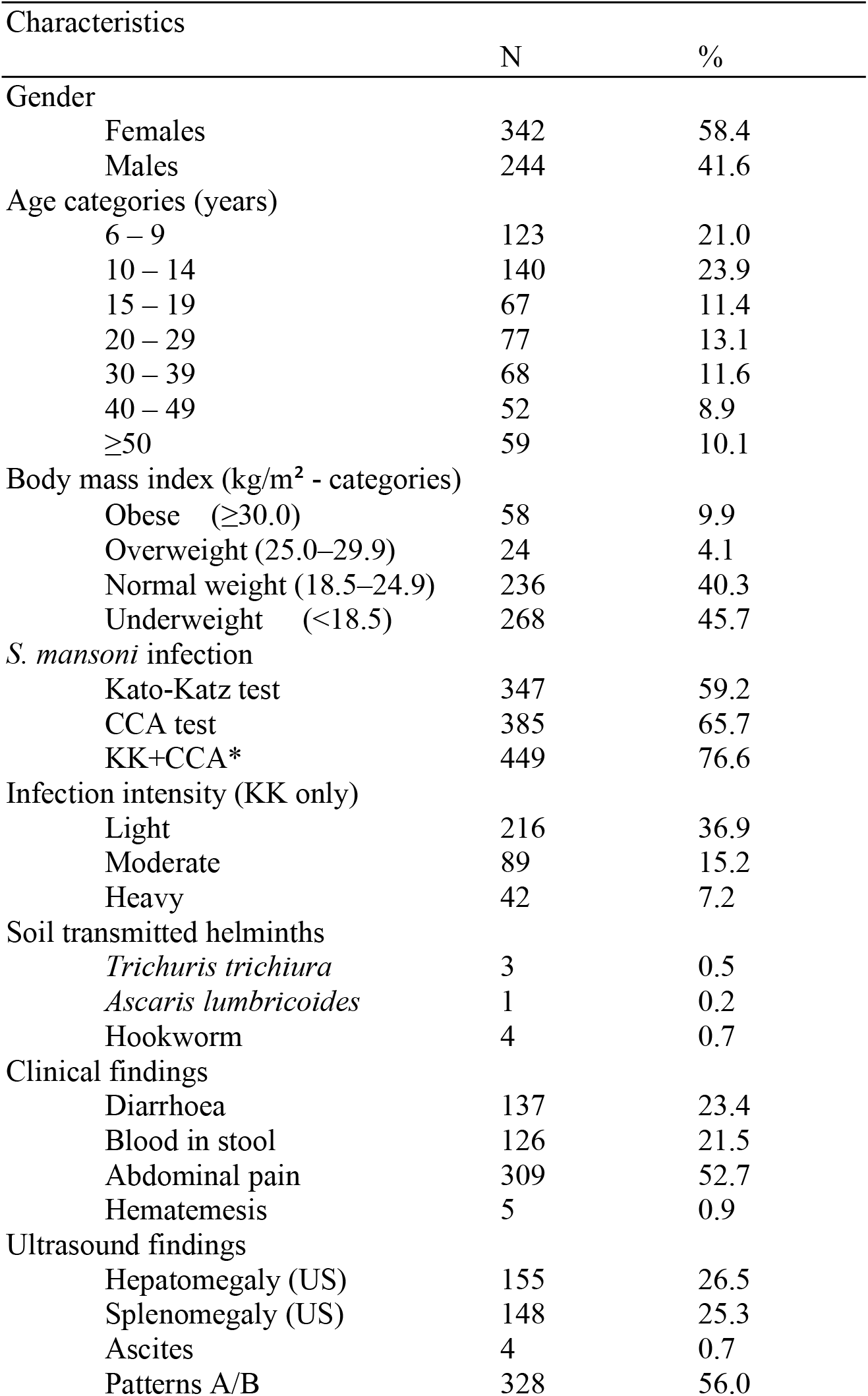
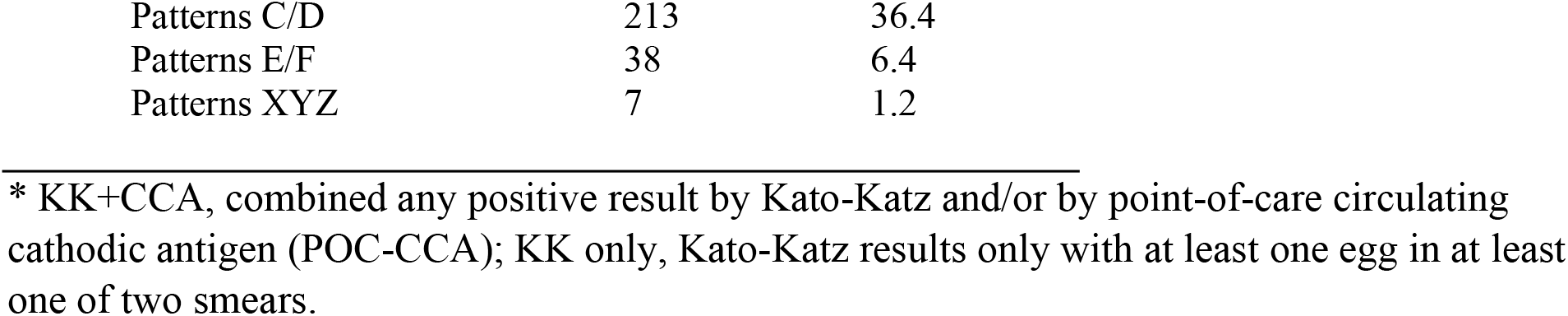
Study sample characteristics in the 2017 Ituri infection and morbidity study. Study conducted in 13 purposively selected villages in Ituri province (n=586).

### Morbidity associated with *S. mansoni* infection

The results of the univariable risk analysis of combined approach are presented in Table 2. Male participants were more likely to be infected with *S. mansoni*, but the increased risk was not statistically significant (OR 1.22, 95% CI 0.82–1.81, *P*=0.318). *S. mansoni* infection was observed more frequently in younger age groups, with prevalence peaking among young adults (Figure 3). Participants aged 50 years and older had a statistically significant reduced risk of infection compared to children aged 6–9 years (Table 2, OR 0.49, 95% CI 0.26–0.92, *P*=0.024).

**Table 2:** Morbidity associated with *S. mansoni* infection in the 2017 study. Results of the univariable analysis of data from 13 purposively selected villages in Ituri province (n=586).

| Characteristics | <u>*S. mansoni* (+)</u> |  | <u>*S. mansoni* (-)</u> |  | OR (95% CI) | p-value |
| --- | --- | --- | --- | --- | --- | --- |
|  | <u>N=449</u> |  | <u>N=137</u> |  |  |  |
|  | n | % | n | % |  |  |
| Gender* |  |  |  |  |  |  |
| Females | 257 | 57.2 | 85 | 62.0 | 1.0 |  |
| Males | 192 | 42.8 | 52 | 38.0 | 1.22 (0.82–1.81) | 0.318 |
| Age categories (years)* |  |  |  |  |  |  |
| 6 – 9 | 92 | 20.5 | 31 | 22.6 | 1.0 |  |
| 10 – 14 | 119 | 26.5 | 21 | 15.3 | 1.61 (0.94–2.76) | 0.078 |
| 15 – 19 | 54 | 12.0 | 13 | 9.5 | 1.10 (0.58–2.07) | 0.769 |
| 20 – 29 | 63 | 14.0 | 14 | 10.2 | 1.53 (0.81–2.89) | 0.184 |
| 30 – 39 | 48 | 10.7 | 20 | 14.6 | 0.77 (0.42–1.42) | 0.396 |
| 40 – 49 | 35 | 7.8 | 17 | 12.4 | 0.73 (0.38–1.43) | 0.359 |
| ≥50 | 38 | 8.5 | 21 | 15.3 | 0.49 (0.26–0.92) | 0.024 |
| STH |  |  |  |  |  |  |
| *T. trichiura* (Y/N)* | 1 | 0.2 | 2 | 1.5 | 0.15 (0.01–1.69) | 0.076 |
| *A. lumbricoides* (Y/N) | 1 | 0.2 | 0 | 0.0 | na |  |
| Hookworm (Y/N)* | 4 | 0.2 | 3 | 2.2 | 0.10 (0.01–0.98) | 0.015 |
| Anthropometry (BMI)* |  |  |  |  |  |  |
| Obese (Y/N) | 36 | 8.0 | 22 | 16.1 | 1.0 |  |
| Overweight (Y/N) | 18 | 4.0 | 6 | 4.4 | 1.83 (0.62–5.40) | 0.264 |
| Normal weight (Y/N) | 183 | 40.8 | 53 | 38.7 | 2.11 (1.14–3.92) | 0.016 |
| Underweight (Y/N) | 212 | 47.2 | 56 | 40.9 | 2.31 (1.25–4.28) | 0.006 |
| Clinical findings |  |  |  |  |  |  |
| Diarrhoea (Y/N)* | 114 | 25.4 | 23 | 16.8 | 1.69 (1.03–2.78) | 0.038 |
| Blood in stool (Y/N) | 99 | 22.1 | 27 | 19.7 | 1.15 (0.72–1.86) | 0.560 |
| Abdominal pain (Y/N) | 238 | 53.0 | 71 | 51.8 | 1.05 (0.72–1.54) | 0.808 |
| Hematemesis (Y/N) | 4 | 0.9 | 1 | 0.7 | 1.22 (0.14–11.05) | 0.858 |
| Ultrasound findings |  |  |  |  |  |  |
| Hepatomegaly (Y/N)* | 125 | 27.8 | 30 | 21.9 | 1.38 (0.87–2.17) | 0.168 |
| Splenomegaly (Y/N)* | 118 | 26.3 | 30 | 21.9 | 1.27 (0.81–2.01) | 0.302 |
| Ascites (Y/N)* | 2 | 0.5 | 2 | 1.5 | 0.30 (0.04–2.17) | 0.207 |
| Pattern A/B (Y/N) * | 246 | 54.8 | 82 | 59.9 | 1.0 |  |
| Pattern C/D (Y/N) | 167 | 37.2 | 46 | 33.6 | 1.21 (0.80–1.83) | 0.363 |
| Pattern E/F (Y/N) | 30 | 6.7 | 8 | 5.8 | 1.25 (0.55–2.84) | 0.593 |
| Pattern XYZ (Y/N) | 6 | 1.3 | 1 | 0.7 | 2.00 (0.24–16.94) | 0.517 |

\* Included in the multivariable analysis. BMI, body mass index (only taken as continuous variable); na, not applicable; Pattern A: normal; Pattern B: “starry sky”; Pattern C: “rings and pipe-stems”; Pattern D “highly echogenic ruff around portal bifurcation”; Pattern E “highly echogenic patches”; Pattern F: “highly echogenic bands and streaks – bird’s claw”; Patterns X, Y, Z indicate pathology different from periportal fibrosis [21].

Intestinal helminth co-infections were negatively associated with *S. mansoni* infection status among them the association with hookworm infection was statistically significant (OR 0.10, 95% CI 0.01–0.98, P=0.015). Study participants who reported an episode of diarrhoea within the preceding two weeks had an increased risk of being infected with *S. mansoni* (OR 1.69, 95% CI 1.03–2.78, *P*=0.038).

Diagnosed hepatomegaly (OR 1.38, 95% CI 0.87–2.17, *P*=0.168), splenomegaly (OR 1.27, 95% CI 0.81–2.01, *P*=0.302) and E/F liver parenchyma patterns (OR 1.25, 95% CI 0.55–2.84, *P*=0.593) were positively but not significantly associated with an *S. mansoni* infection.

The univariable risk analysis for Kato-Katz and POC-CCA diagnosis are given in supplementary Table S1 and S3, respectively. They provided a very similar risk pattern. Noteworthy is that when results of Kato-Katz diagnosis alone are considered male participants had a significant higher risk for a *S. mansoni* infection (Table S1, OR 1.44, 95% CI 1.03–2.03, *P*=0.033) and hepatomegaly (Table S1, OR 1.41, 95% CI 0.96–2.06, *P*=0.079) was not significantly associated with *S. mansoni* infection, whereas splenomegaly (Table S1, OR 1.61, 95% CI 1.09–2.39, *P*=0.017) was significantly associated with *S. mansoni* infection.

**Figure 2:** *S. mansoni* infection prevalence by age in the 2017 Ituri province morbidity study (n=586).

Overall (green-solide line), female (red-dash line), and male (maroon-long-dash line).

The age distribution of reported diarrhoea and blood in stool, as well as the ultrasonographically assessed hepato- and splenomegaly displayed an age distribution resembling the *S. mansoni* infection with peaks in the adolescent and adult age groups (Figure 3).

**Figure 3:**
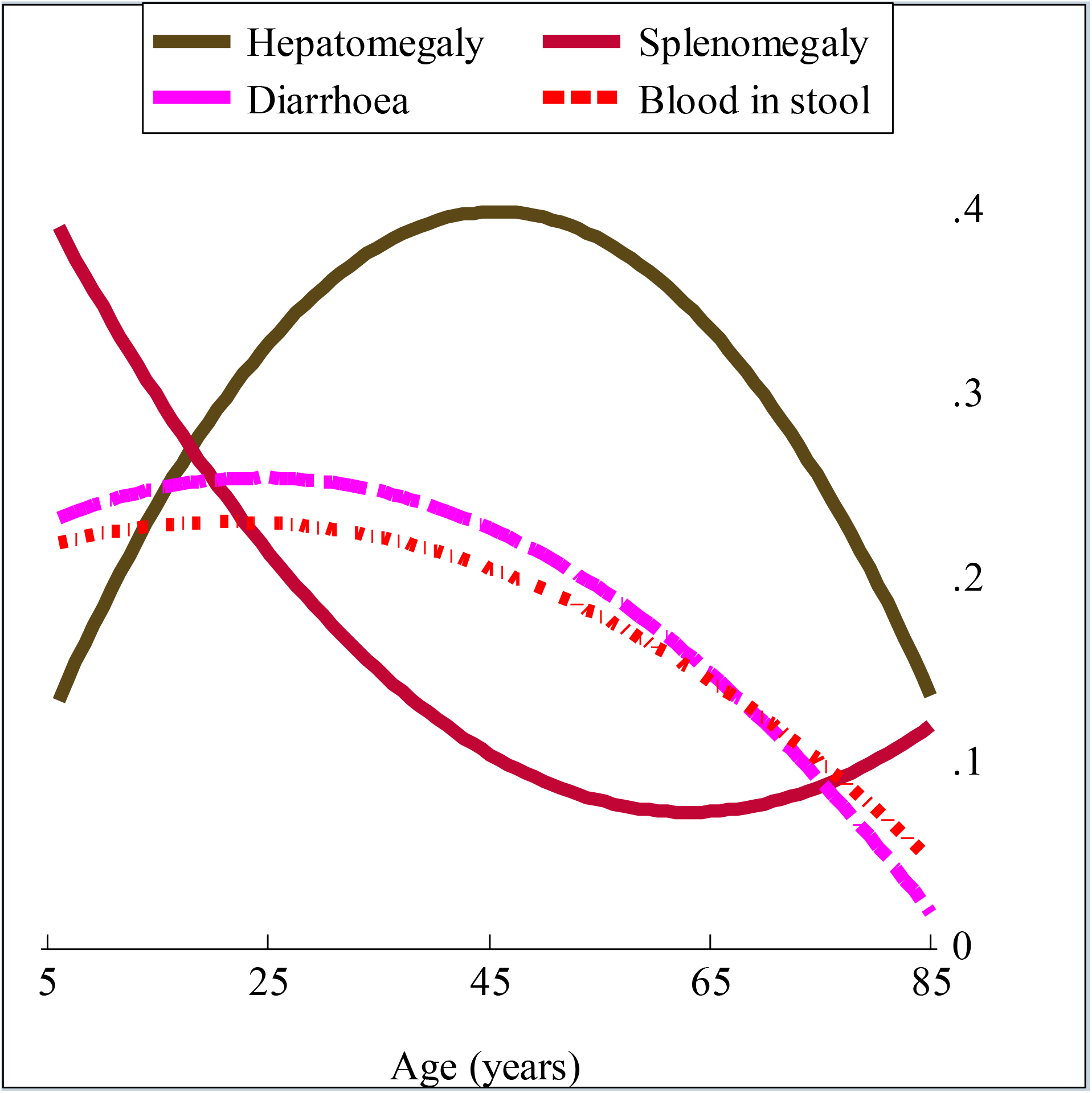
Age distribution of intestinal and hepatosplenic morbidity in the 2017 Ituri province morbidity study (n=586). Hepatomegaly (olive-solide line), splenomegaly (cranberry-solide line), diarrhoea (magenta-long-dash line), and blood in stool (red-short-dash-dot line).

The risk analysis took in account results of the combined Kato-Katz and POC-CCA diagnostic approach (Table 3). These results were broadly consistent when using Kato-Katz or POC-CCA alone. However, there were some discrepancies. First, intestinal morbidity indicator, such as diarrhoea, was significantly associated with an *S. mansoni* infection (OR 1.69, 95% CI 1.03–2.78, *P*=0.038). This is also the case when results of Kato-Katz test alone are taken in account (OR 1.85, 95% CI 1.22–2.79, *P*=0.003). However, using results of POC-CCA alone, the association became not significant (OR 1.42, 95% CI 0.93–2.16, *P*=0.101). Other indicators including the presence of blood in stool, abdominal pain, and history of hematemesis were not associated regardless of the diagnostic approaches used. Second, hepatomegaly was not significantly associated with an *S. mansoni* infection using the combined (OR 1.38, 95% CI 0.87–2.17, *P*=0.168), even using POC-CCA (OR 1.33, 95% CI 0.89–1.98, *P*=0.158), or Kato-Katz test alone (OR 1.41, 95% CI 0.96–2.06, *P*=0.079). Splenomegaly was significantly associated with an *S. mansoni* infection using Kato-Katz test alone (OR 1.61, 95% CI 1.09–2.39, *P*=0.017). The association was not significant when using the combined and POC-CCA diagnostic approaches (OR 1.27, 95% CI 0.81–2.01, *P*=0.302) and (OR 1.17, 95% CI 0.78–1.74, *P*=0.451), respectively. Third, an abnormal liver parenchyma pathology (combined patterns E/F) was significantly associated with *S. mansoni* infection when using either Kato-Katz test alone (OR 2.25, 95% CI 1.05–4.80, *P*=0.032). The association was not significant when using POC-CCA alone or the combined approaches (OR 1.20, 95% CI 0.58–2.47, *P*=0.618) and (OR 1.25, 95% CI 0.55–2.84, *P*=0.593), respectively.

Ten variables were included in the multivariable logistic regression analysis, the results of which are displayed in Table 3. Age was negatively associated with *S. mansoni* infection (adjusted odds ratio [aOR] 0.98; 95% CI 0.96–0.99, *P*=<0.001) while gender showed not significantly associated with the *S. mansoni* infection (aOR 1.15; 95% CI 0.74–1.79, *P*=0.524).

Of the morbidity indicators investigated, diarrhoea (aOR 1.69; 95% CI 0.99–2.89, *P*=0.053) and hepatomegaly (aOR 1.58; 95% CI 0.96–2.61, *P*=0.071) were associated with *S. mansoni* infection with a borderline significance level. Patients with abnormal liver parenchyma patterns (aOR 1.13; 95% CI 0.98–1.31, *P*=0.100) did not have an increased risk for *S. mansoni* infection.

**Table 3.** Morbidity associated with *S. mansoni* infection in the 2017 study based on Kato-Katz and POC-CCA diagnostic approach. Results of the multivariable analysis of data from 13 purposively selected villages in Ituri province (n=586).

| Risk factors | aOR (95% CI) | Std. Err. | z | p-value |
| --- | --- | --- | --- | --- |
| Demographic risk factors |  |  |  |  |
| Age | 0.98 (0.96–0.99) | 0.006 | -3.64 | <0.001 |
| Gender (Male/Female) | 1.15 (0.74–1.79) | 0.259 | 0.64 | 0.524 |
| Anthropometric risk factors |  |  |  |  |
| BMI | 1.00 (0.95–1.06) | 0.027 | 0.15 | 0.878 |
| Clinical finding |  |  |  |  |
| Diarrhoea | 1.69 (0.99–2.89) | 0.461 | 1.94 | 0.053 |
| Blood in stool | 0.90 (0.54–1.50) | 0.237 | -0.41 | 0.683 |
| Ultrasound findings |  |  |  |  |
| Hepatomegaly (Yes/No) | 1.58 (0.96–2.61) | 0.404 | 1.80 | 0.071 |
| Splenomegaly (Yes/No) | 0.91 (0.55–1.50) | 0.230 | -0.37 | 0.712 |
| Ascites (Yes/No) | 0.21 (0.03–1.69) | 0.225 | -1.46 | 0.144 |
| Liver pathology (Yes/No) | 1.13 (0.98–1.31) | 0.085 | 1.65 | 0.100 |
| Co-infection |  |  |  |  |
| Hookworm (Yes/No) | 0.08 (0.01–0.81) | 0.093 | -2.14 | 0.033 |

aOR: adjusted odds ratio in multivariable analysis; CI: confidence interval. BMI, body mass index (only taken as continuous variable).

At the village level, the prevelance of hepatomegaly (Figure 4) and splenomegaly (Figure 5) increased with the prevalence of *S. mansoni* infection. Four patients were diagnosed with ascites; all of whom were residents of villages where overall *S. mansoni* prevalence exceeded 80%.

**Figure 4:**
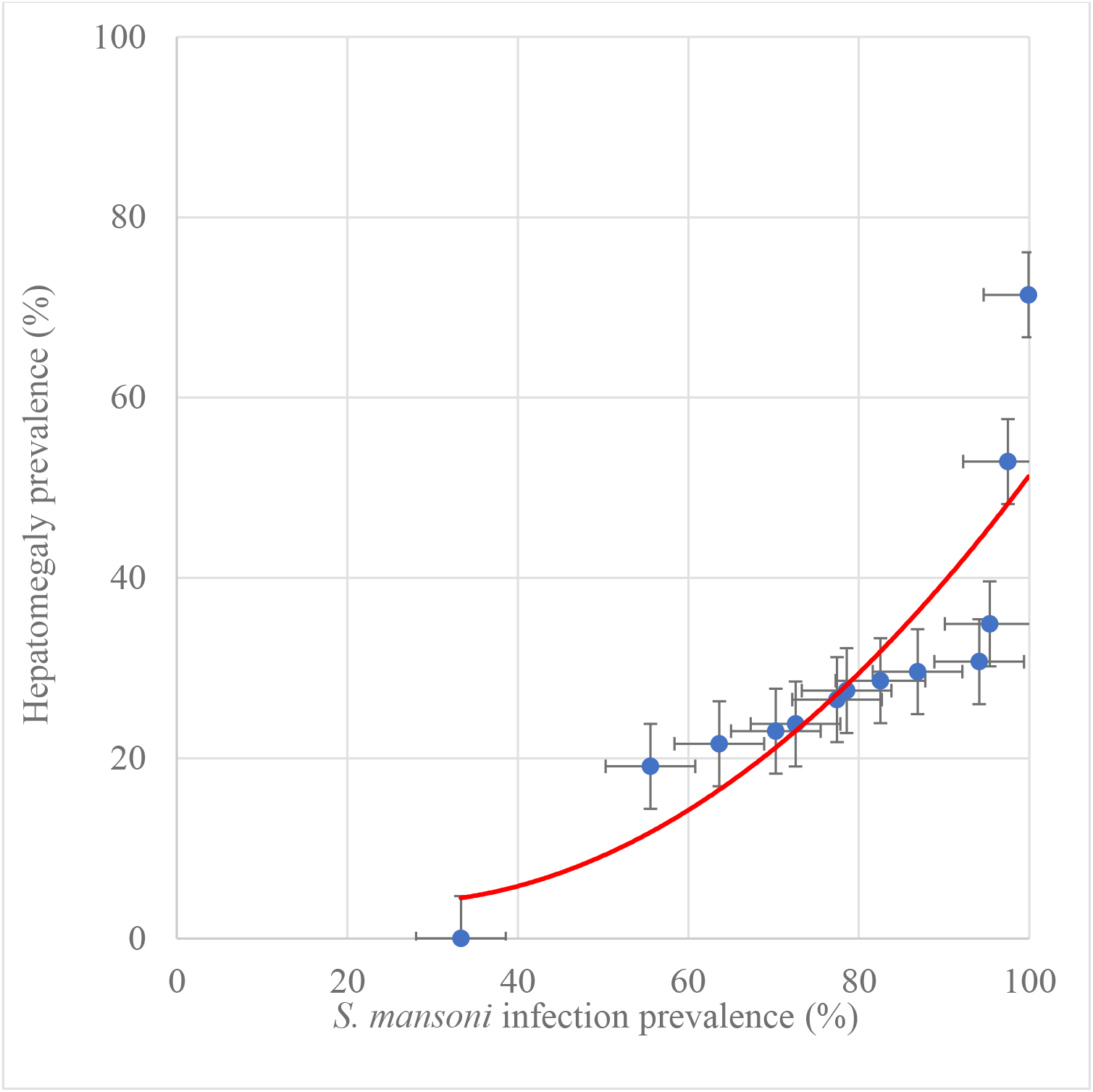
Association of hepatomegaly and *S. mansoni* infection prevalence at village level in the 2017 Ituri morbidity study (n=586).

**Figure 5:**
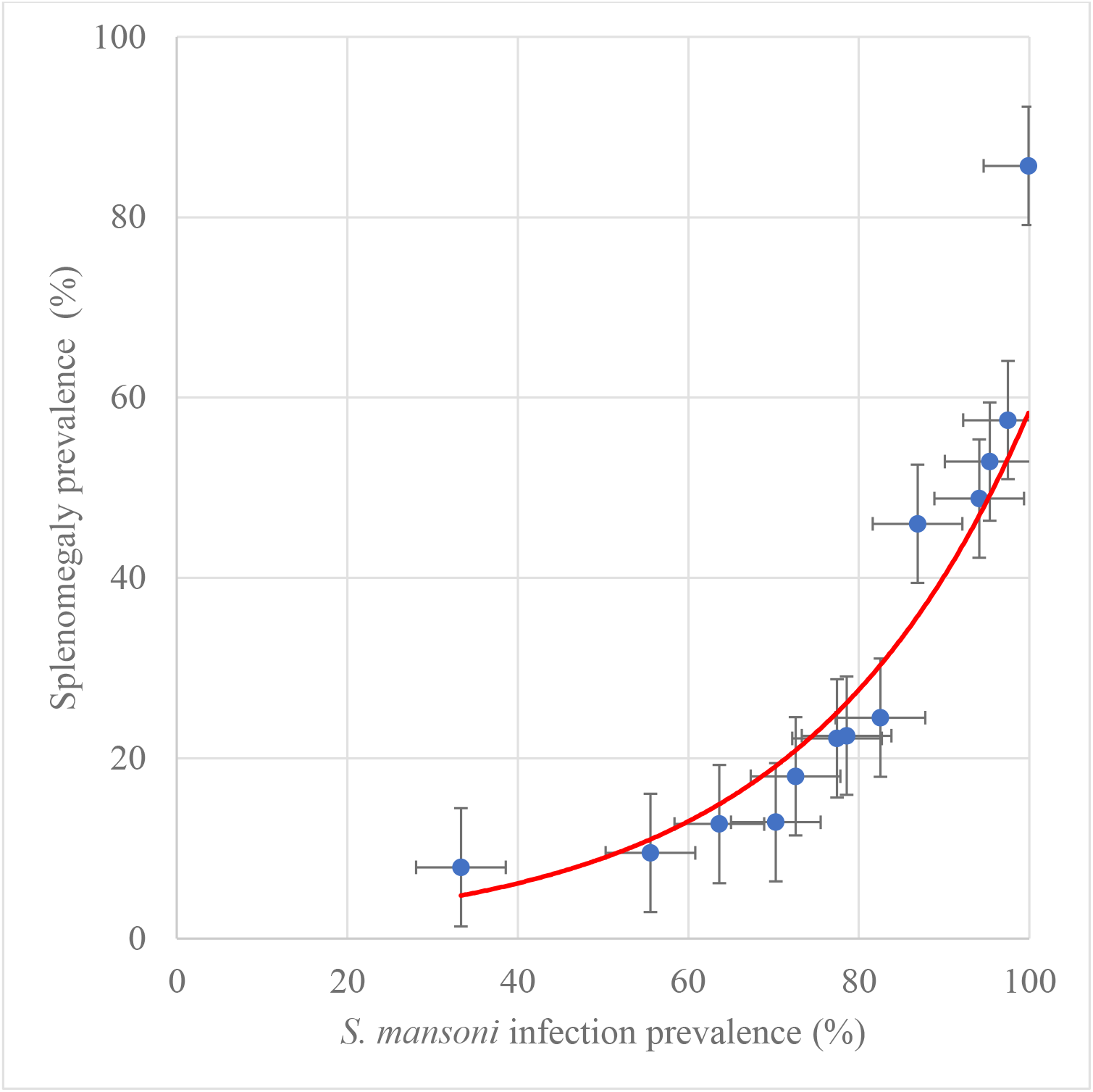
Association of splenomegaly and *S. mansoni* infection prevalence at village level in the 2017 Ituri morbidity study (n=586).

*S. mansoni* infection intensity varied greatly among individuals, with a maximum of 4,497.6 EPG and a mean infection intenstity of 109.7 EPG. Table 4 presents the infection intensity according to gender, age, helminth co-infection and morbidity categories. Infection intensity levels were similar in the two gender groups (*P*=0.198). The age distribution of the infection intensity levels followed the age-infection prevalence curve. Heavy-intensity infections were mostly found (12.9%) among adolescents aged 10–14 years,while no one in the oldest age group (50 years and older) had a heavy-intensity infection (*P*=0006). Heavy-intensity infections were significantly higher amongpatients co-infected with *Ascaris lumbricoides* (13.0%, *P*=0.005) and underweight participants (10.1%, *P*=0.033).

There was a significantly higher prevalence of reported diarrhoea (40.5%, *P*=0.004) and blood in the stool (52.4%, P<0.001) among patients in the heavy-intensity infection group compared to the other infection intensity groups.

Among patients with heavy-intensity infections, the prevalence of splenomegaly (57.1%) was significantly higher than among other infection intensity groups (P<0.001), while the prevalence of hepatomegaly (38.1%) was not statistically different compared with the other infection intensity groups (*P*=0.073). When stratified by age, patients with an enlarged liver and/or spleen bore a higher infection intensity burden compared to those with a normal-sized liver and spleen in the same age group (Figure 6 and Table S6). In general, younger patients (children: aged <18 years) experienced more high-intensity infections compared to those in older age groups (adults: aged > 18 years).

**Table 4:** *S. mansoni* infection intensity by morbidity in the 2017 study. Study conducted in 13 purposively selected villages in Ituri province (n=586). Only results of the Kato-Katz test have been considered in this analysis.

| Characteristics | <u>Negative</u> | | <u>*S. mansoni* infection intensity</u> | | | | <u>Heavy</u> | | $\chi^2$ | p-value |
| --- | --- | --- | --- | --- | --- | --- | --- | --- | --- | --- |
|  | n | % | <u>Light</u><br>n | % | <u>Moderate</u><br>n | % | n | % |  |  |
| Overall | 239 | 40.8 | 216 | 36.9 | 89 | 15.2 | 42 | 7.2 |  |  |
| Gender |  |  |  |  |  |  |  |  |  |  |
| Females | 152 | 44.4 | 117 | 34.2 | 50 | 14.6 | 23 | 6.7 |  |  |
| Males | 87 | 35.7 | 99 | 40.6 | 39 | 16.0 | 19 | 7.8 | 4.66 | 0.198 |
| Age categories (years) |  |  |  |  |  |  |  |  |  |  |
| 6 – 9 | 50 | 40.7 | 47 | 38.2 | 18 | 14.6 | 8 | 6.5 |  |  |
| 10 – 14 | 49 | 35.0 | 49 | 35.0 | 24 | 17.1 | 18 | 12.9 |  |  |
| 15 – 19 | 19 | 28.0 | 25 | 37.3 | 17 | 25.4 | 6 | 9.0 |  |  |
| 20 – 29 | 27 | 35.0 | 32 | 41.6 | 11 | 14.3 | 7 | 9.1 |  |  |
| 30 – 39 | 31 | 45.6 | 26 | 38.2 | 9 | 13.3 | 2 | 2.9 |  |  |
| 40 – 49 | 27 | 51.9 | 18 | 34.6 | 6 | 11.6 | 1 | 1.9 |  |  |
| ≥50 | 36 | 61.0 | 19 | 32.2 | 4 | 6.8 | 0 | 0.0 | 36.68 | 0.006 |
| Soil-transmitted helminths |  |  |  |  |  |  |  |  |  |  |
| *T. trichiura* (Y/N) | 2 | 0.8 | 1 | 0.5 | 0 | 0.0 | 0 | 0.0 | 1.18 | 0.758 |
| *Ascaris* (Y/N) | 0 | 0.0 | 0 | 0.0 | 0 | 0.0 | 1 | 2.4 | 13.0 | 0.005 |
| Hookworm (Y/N) | 3 | 1.3 | 1 | 0.5 | 0 | 0.0 | 0 | 0.0 | 2.21 | 0.530 |
| Anthropometry (BMI) |  |  |  |  |  |  |  |  |  |  |
| Obese | 33 | 56.9 | 20 | 34.5 | 4 | 6.9 | 1 | 1.7 |  |  |
| Overweight | 13 | 54.2 | 9 | 37.5 | 2 | 8.3 | 0 | 0.0 |  |  |
| Normal weight | 96 | 40.7 | 89 | 37.7 | 37 | 15.7 | 14 | 5.9 |  |  |
| Underweight | 97 | 36.2 | 98 | 36.5 | 46 | 17.2 | 27 | 10.1 | 18.15 | 0.033 |
| Clinical findings |  |  |  |  |  |  |  |  |  |  |
| Diarrhoea (Y/N) | 41 | 17.2 | 56 | 25.9 | 23 | 25.8 | 17 | 40.5 | 13.11 | 0.004 |
| Blood in stool (Y/N) | 42 | 17.6 | 32 | 14.8 | 30 | 33.7 | 22 | 52.4 | 39.49 | <0.001 |
| Abdom. pain (Y/N) | 124 | 51.9 | 107 | 49.5 | 52 | 58.4 | 26 | 61.9 | 3.53 | 0.317 |
| Hematemesis (Y/N) | 3 | 1.3 | 1 | 0.5 | 1 | 1.1 | 0 | 0.0 | 1.28 | 0.733 |
| Ultrasound findings |  |  |  |  |  |  |  |  |  |  |
| Hepatomegaly (Y/N) | 54 | 22.6 | 65 | 30.1 | 20 | 22.5 | 16 | 38.1 | 6.95 | 0.073 |
| Splenomegaly (Y/N) | 48 | 20.1 | 48 | 22.2 | 28 | 31.5 | 24 | 57.1 | 28.88 | <0.001 |
| Ascites (Y/N) | 3 | 1.3 | 0 | 0.0 | 1 | 1.1 | 0 | 0.0 | 3.19 | 0.364 |
| Pattern A/B (Y/N) | 146 | 44.5 | 120 | 36.6 | 43 | 13.1 | 19 | 5.8 |  |  |
| Pattern C/D (Y/N) | 80 | 37.6 | 79 | 37.1 | 37 | 17.4 | 17 | 8.0 |  |  |
| Pattern E/F (Y/N) | 10 | 26.3 | 13 | 34.2 | 9 | 23.7 | 6 | 15.8 |  |  |
| Pattern XYZ (Y/N) | 3 | 42.9 | 4 | 57.1 | 0 | 0.0 | 0 | 0.0 | 14.46 | 0.107 |

BMI, body mass index; Abdom.: abdominal. Pattern A: normal; Pattern B: “starry sky”; Pattern C: “rings and pipe-stems”; Pattern D “highly echogenic ruff around portal bifurcation”; Pattern E “highly echogenic patches”; Pattern F: “highly echogenic bands and streaks – bird’s claw”; Patterns X, Y, Z indicate pathology different from periportal fibrosis. [21].

*S. mansoni* infection intensity varied considerably among patients with different liver parenchyma pathologies (Tables S5 and S7). In general, more heavy-intensity infections were observed among patients with more severe liver morbidity patterns. That is, the number of individuals with heavy-intensity infections increased with the severity of the liver parenchyma pattern, from normal liver parenchyma patterns A and B (5.8%) to the most severe “bird’s claw” patterns E and F (15.8%). The association was not statistically significant (*P*=0.107). When stratified by age, a clear association emerged between increased number of high-intensity infections and increasingly abnormal liver pathologies (Figure 7). Liver parenchyma worsened, from A and B normal patterns to C and D mild PPF patterns, and to E and F severe PPF patterns, the median infection intensity increased. However, taken alone, patients with pattern F had similar-or lower-intensity infections than patients with less severe morbidity patterns (Table S7).

**Figure 6:**
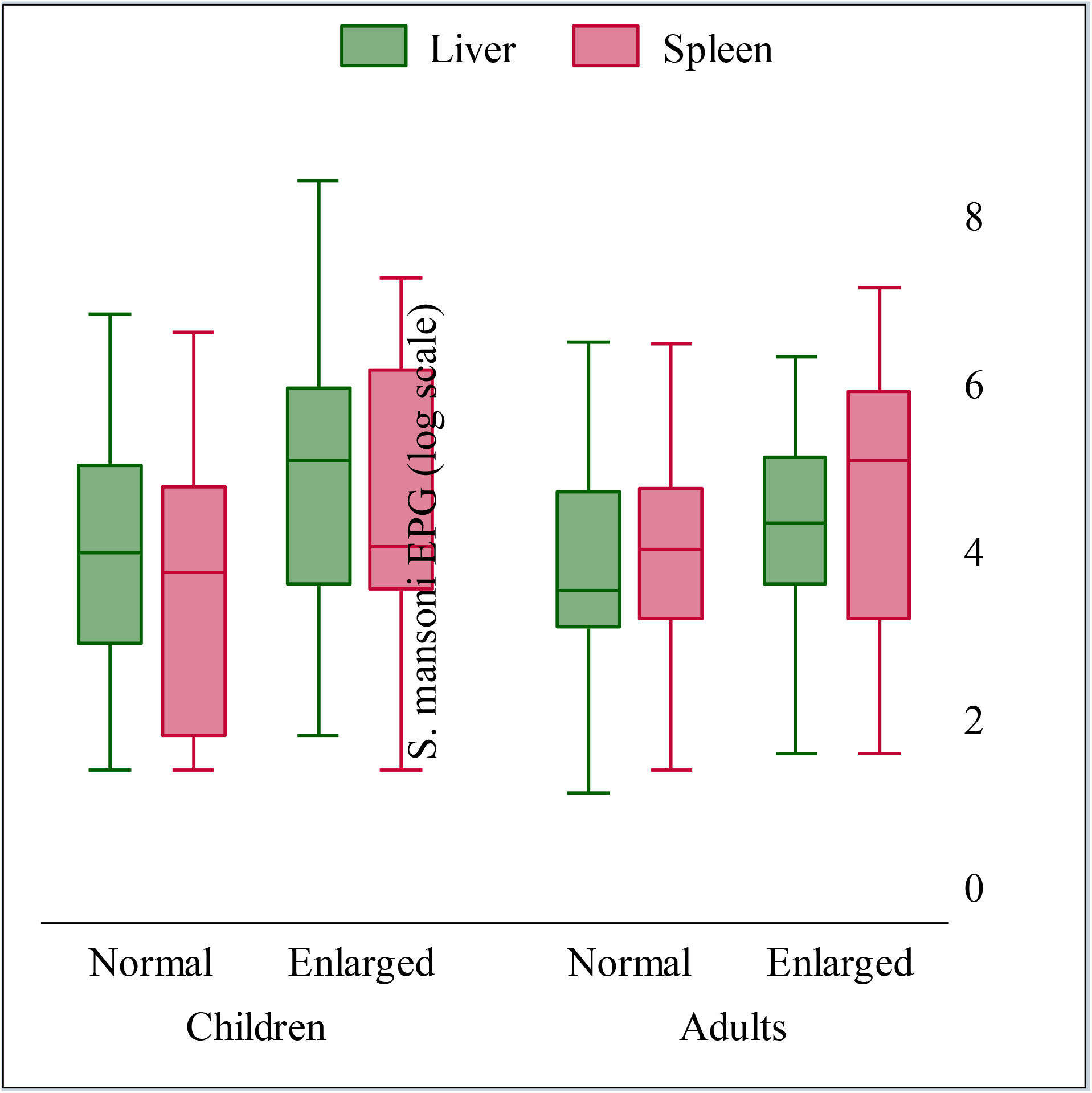
*S. mansoni* infection intensity by hepatomegaly and splenomegaly and age in the 2017 Ituri morbidity study (n=586). Hepatomegaly (green) and splenomegaly (cranberry).

**Figure 7:**
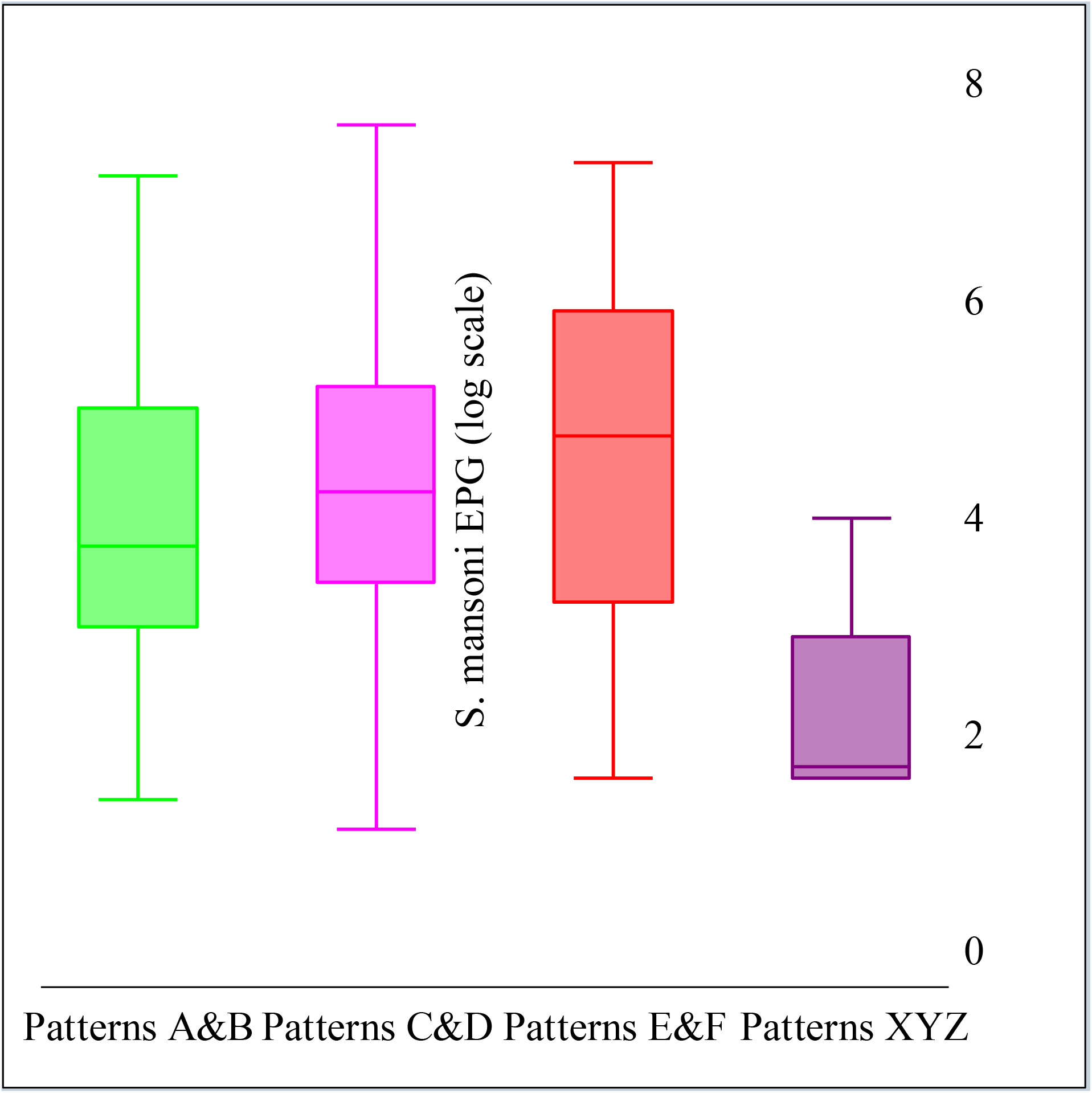
*S. mansoni* infection intensity by liver parenchyma patterns and age in the 2017 Ituri morbidity study (n=586). Patterns A&B: normal (lime); Patterns C/D: mild PPF (magenta); Pattern E/F: severe PPF (red); Pattern XYZ: other no PPF pathologies (purple).

## Discussion

The focus of the World Health Organization’s roadmap for neglected tropical diseases 2012–2030 with regard to schistosomiasis is control of morbidity. This is especially the case in the African region where transmission levels in several countries, including the Democratic Republic of the Congo are high, and elimination is not yet scheduled. The key recommended strategic approach is preventive chemotherapy with praziquantel. Mass treatment of population exposed to risk of infection, at regular intervals, prevents the development of high-intensity infections and hence, of morbidity [23, 24].

Our study provides, for the first time, comprehensive baseline data showing a high intestinal and hepatosplenic morbidity burden in Ituri province; a burden that is associated with *S. mansoni* infection at both the individual and community level. At present, we cannot quantify the extent to which *S. mansoni* infection alone is responsible for the observed morbidity (or whether other infections contribute). To answer this question, additional investigations are needed. However, the high degree of intestinal and hepatosplenic morbidity present in Ituri province warrants immediate and comprehensive control activities.

Although minimizing morbidity is the target of schistosomiasis control efforts, control programmes rarely collect (baseline) and monitor morbidity data. Instead, they largely rely on monitoring infection intensities, which are linked to morbidity and much easier to assess than intestinal and hepatosplenic morbidity. Consequently, little is known about the morbidity burden of schistosomiasis [6].

Control programmes have been conducted successfully in colonial time, even with means that today may be considered outdated. However, with these means the disease was well controlled [15]. Since the independence of the country, no large-scale activity aimed at combating the disease has ever been undertaken until 2012. Since this date, a public control program targeting school children started in the country and it was launched in the Ituri province in 2016.

In this study, we assessed the magnitude of the intestinal and hepatosplenic morbidity burden in Ituri province, DRC, and investigated its associations with *S. mansoni* infection status. To that end, we conducted a cross-sectional study in 13 *S. mansoni* endemic villages. We enrolled all household member six years and older and assessed their *S. mansoni* infection status and their intestinal and hepatosplenic morbidity.

We found a high degree of intestinal and hepatosplenic morbidity. About one quarter of the study participants reported diarrhoea (23.4%) and blood in the stool (21.5%). Upon ultrasonography examination, almost one-quarter was diagnosed with hepatomegaly (26.5%); almost two-thirds (25.3%) had splenomegaly, and more than half (42.8%) had abnormal liver parenchyma (pattern C–F). Five patients reported an experience of hematemesis and four patients had ascites.

In our study population, we found a high prevalence of *S. mansoni* infection (76.6%). As POC-CCA does not provide information on infection intensity, the analysis of infection intensity was done based on the Kato-Katz results only. Light-, moderate- and heavy-intensity infections were diagnosed in high frequencies of 36.9%, 15.2% and 7.2%, respectively. Only few cases of soil-transmitted helminths with an expected very small impact on the morbidity were diagnosed during this study. *S. mansoni* infection prevalence and intensity was highest in the adolescent and young adult age groups. The prevalence of intestinal and hepatosplenic morbidity indicators showed a very similar age distribution (although hepato- and splenomegaly peaked in older age groups), and at village level, hepatosplenic morbidity prevalence increased with infection prevalence. Both observations suggesting a close link between morbidity and *S. mansoni* infection. Furthermore, at individual level, we found an increased risk of hepatomegaly and splenomegaly in *S. mansoni-*infected patients, confirming the association found at village level and the similarly shaped age distributions. The findings are also consistent with documented hepatosplenic morbidity associated with *S. mansoni* infection [25, 26]. Hence, providing further evidence that *S. mansoni* infection is a major contributor to the overall observed morbidity.

We found three notable differences in the risk results when relying on the diagnostic results from the Kato-Katz technique only. However, as we needed to get the respective advantages of both more specific (Kato-Katz) and more sensitive (POC-CCA) tests, we considered the results of the Kato-Katz + POC-CCA combined approach. First, reported diarrhoea was significantly associated with *S. mansoni* infection; second, pathological changes of the liver parenchyma were associated with *S. mansoni* infection as was hepatomegaly; and, third, splenomegaly was not associated with *S. mansoni* infection. Using the Kato-Katz technique alone to diagnose *S. mansoni* infection reduces the overall sensitivity of the diagnostic approach due to the low sensitivity of the technique itself [27, 28]. Hence, on average, those diagnosed with an *S. mansoni* infection are more likely to have a higher infection intensity in comparison to the combined diagnostic approach. From these observations, we see that subtle morbidity increases — such as reported diarrhoea and pathological changes in the liver parenchyma — become statistically significant. Indeed, for both morbidity indicators, we observed an association with *S. mansoni* infection intensity. Patients with diarrhoea had the highest prevalence of heavy-intensity infections (Table 2) and those with abnormal liver parenchyma patterns E-F had the highest mean infection intensities (Figure 4 and Table S7). However, the importance of the association of parasitological diagnosis with morbidity was much more at the community (village) level than at the individual level.

In our study, we found that patients with abnormal liver parenchyma pattern F displayed lower *S. mansoni*s infection intensity and risk compared to the E pattern. The highly echogenic bands and streaks corresponding to the pattern F extend from main portal vein and its bifurcation to the liver surface. Most habitually, this pattern is manifested in very advanced cases of liver fibrosis and is frequently conglomerated with other changes [21]. Patients who displayed this pattern were often older, thus, they have little exposure to water contact, and some have been already treated once, twice or more times with praziquantel. Consequently, they may be free of infection or may have light infection intensities. It is also known that treatment with praziquantel could not halt progression of organ damage in some individuals. This situation may be due to immunological and genetic factors as well as other factors, including malaria, viral hepatitis and/or concomitant alcohol consumption [5, 29–31].

Quantifying schistosomiasis morbidity is a challenging [6, 32] and controversial matter [4]. Morbidity associated with *Schistosoma* infection is unspecific. Hence, the observed morbidity pattern might be provoked partially by or in combination with other pathogens, such as other helminth species, protozoa, bacteria and viruses. Given that multiple infections are frequent in tropical Africa, a combination of infections is most likely responsible for the observed morbidity. In Ituri province, other parasitic infections, such as malaria (i.e. *Plasmodium falciparum)*, and other infections with hepatosplenic affinity, such as viral hepatitis, are prevalent [3, 5, 33, 34] and may have contributed to the hepatosplenic morbidity pattern observed. The time gap between infection and the occurrence of measurable morbidity further complicates efforts to assess the association between infection and morbidity. Furthermore, organomegaly is sometimes described as normally present in children; it then regresses and disappears in adulthood [5, 35–37].

Nevertheless, these findings cannot in any way reduce the value of abdominal ultrasound in the diagnosis of liver pathologies associated with *S. mansoni* infection. However, ultrasonography devices are rarely available in the poor-resource settings. In Ituri province, the device may be found only in the provincial hospital and in some district hospitals and private clinics. In our field work, we found that the use of portable devices is feasible in the villages. However, in the remotest rural areas, the usage is challenging as electricity was not available. It requires additional equipment such as solar panels and rechargeable batteries. Despite of these shortcomings, abdominal ultrasound adds crucial information on the liver morbidity in the community and hence, indispensable information on the public health burden of *S. mansoni* infection [38]. Furthermore, severe cases are likely to be early diagnosed and adequately addressed.

In our study, we encountered patients with severe complications from *S. mansoni* infection, which further underscore the importance of the infection’s morbidity burden. Four people (0.9%) reported a history of hematemesis and two people (0.5%) with ascites. The finding appears to corroborate the health service’s statistics report from the Angumu health district (on the shore of Lake Albert), which declares that hematemesis is a frequent medical emergency and that adults have died after vomiting blood in this area. Oesophageal varices remain silent until they rupture and irreversible damage occurs [34]. Angumu health district is a remote area and well known for its high blood transfusion rates. Patients vomiting blood often reach the hospital too late, leading to the worst medical outcome.

The morbidity levels we observed are consistent with those measured by Ongom and Bradley [18], who found serious morbidity, including diarrhoea and abdominal pain, in a schistosomiasis endemic community on Lake Albert in Uganda. Other studies of *S. mansoni* endemic communities outside of the DRC present similar morbidity levels [37–39]. Very few studies of morbidity due to schistosome infection in DRC exist [34, 40]. Our study contributes to the country’s knowledge base and may offer a baseline for future intervention studies to determine the exact extent of morbidity associated with *S. mansoni* infection.

Our study, as many others, presents some limitations. We conducted our study in purposively sampled villages known to have a high prevalence of *S. mansoni*. Thus, the examined population is not representative of the entire province but rather of high transmission areas. Furthermore, ongoing civil unrest in Ituri province creates a challenging security situation, which only afforded us a short time in each village. For this reason, only one stool sample could be collected from each study participant. Finally, limited available resources did not allow us to examine participants for parasitic, bacterial and viral co-infections, which could have helped to better explain the degree of morbidity linked to *S. mansoni* infection.

Other limitations include the lack of the diagnosis of blood coinfections and the definition of the body mass index (BMI) cut-off that did not take in account variations among the study population as it is recommended [41]. Concerning the first limitation, we needed to exclude as much as possible invasive procedures and vulnerable individuals. Indeed, blood sampling is very sensitive in our study area as population is reluctant to blood drawing. It would have required a lot of effort and time for the study. In addition, the compliance of the study participants would have been greatly reduced. Hence, coinfections such as malaria [42], viral hepatitis [43], human immune virus (HIV) as well as other similar diseases endemic in the province were not diagnosed. However, despite not having excluded these confounding factors, the association of S. mansoni infection with hepatosplenic and intestinal morbidity remains highly significant.

As for the BMI cut-off values, we randomly defined in four categories including underweight (<18.5 kg/m^2^), normal weight (18.5–24.9 kg/m^2^), overweight (25.0–29.9 kg/m^2^), and obese (≥30 kg/m^2^) population. This random clustering could be likely to introduce a selection bias in our results. However, in the risk analysis, we used the BMI as a continuous variable instead of the categorical one.

## Conclusion

Schistosomiasis mansoni is an important public health problem in the Ituri province and an appropriate control is not yet being carried. The existing public programme for the control of the disease was launched in 2016 in the province. It is based on preventive chemotherapy and aims at controlling infection among school children. Our results show that both infection and the related morbidity are very high in the province. This situation calls to most vigorous and efficient control measures to correctly address this scourge.

## Data Availability

Data available.

## Acknowledgements

We are grateful to all the study participants both in 2016 and in 2017. Our sincere thanks to the research teams in this two-years period. We thank the Provincial Health Division officers, all the Health Districts officers, and the Institut Supérieur des Techniques Médicales de Nyankunde staff for their support and the local authorities for their support during the fieldwork.

## Supplementary information captions

**Tables**

**Table S1: Univariable associations with *S. mansoni* infection in the 2017 morbidity study**.

Study conducted in 13 purposively selected villages in Ituri province (n=586). Only diagnostic results of Kato-Katz (KK) test have been considered.

Table S2: Risk factors for morbidity due to *Schistosoma mansoni* infection, 2017 study.

Results of the multivariable analysis of risk factors for morbidity due to *Schistosoma mansoni infection among participants from 13 villages in Ituri province (n=586)*. Only diagnostic results of Kato-Katz (KK) test have been considered.

**Table S3: Univariable associations with *S. mansoni* infection in the 2017 morbidity study**.

Study conducted in 13 purposively selected villages in Ituri province (n=586). Only diagnostic results of the point-of-care circulating cathodic antigen (POC-CCA) test have been considered.

**Table S4: Risk factors for morbidity due to *Schistosoma mansoni* infection, 2017 study**.

Results of the multivariable analysis of risk factors for morbidity due to *Schistosoma mansoni infection among participants from 13 villages in Ituri province (n=586)*. Only diagnostic results of the point-of-care circulating cathodic antigen (POC-CCA) test have been considered.

**Table S5: Prevalence of periportal fibrosis (PPF) by age, sex, village, and *S. mansoni* infection status in the 2017 study**.

Results from 13 purposively selected villages of Ituri province (n=586). Prevalence with combined approach and Kato-Katz based intensity.

**Table S6: Prevalence of hepatomegaly, splenomegaly and overall organomegaly by age, sex, village, and *S. mansoni* infection status in the 2017 study**.

Results from 13 purposively selected villages of Ituri province (n=586). Prevalence with combined approach.

**Table S7: *S. mansoni* infection intensity by liver patterns in the 2017 study**.

Study conducted in 13 purposively selected villages in Ituri province (n=586). Only results of the Kato-Katz test have been considered in this analysis.

**Table S8: Univariable analysis of risk of liver patterns and periportal fibrosis (PPF) by *S. mansoni* infection status with different diagnostic approaches in the 2017 study**.

Results from 13 purposively selected villages of Ituri province (n=586). Analysis with Kato-Katz alone, POC-CCA alone, and with the combined approach of Kato-Katz+POC-CCA.

Figures

**Figure S1:** Liver image patterns associated with schistosomiasis, by [21].

